# Change targets, messaging, and content delivery for a community-engaged social media campaign addressing HIV-related stigma in Peru: a qualitative study

**DOI:** 10.1101/2025.02.12.25321801

**Authors:** Alyson Nunez, Marguerite Curtis, Milagros Wong, Kristin A. Kosyluk, Jerome T. Galea, Molly F. Franke, Renato A. Errea

**Affiliations:** Department of Global Health and Social Medicine, Harvard Medical School, Boston, Massachusetts, USA; Socios En Salud Sucursal Perú, Lima, Peru; Department of Mental Health Law and Policy, University of South Florida, Tampa, Florida, USA; School of Social Work, University of South Florida, Florida, USA

**Author notes:** Corresponding author: Molly Franke, 641 Huntington Avenue, Boston, Massachusetts 02115, United States. These authors have contributed equally to the work (Alyson Nunez, Marguerite Curtis). These authors have contributed equally to the work (Molly F. Franke, Renato A. Errea).

**Keywords:** discrimination, youth, adolescents, men who have sex with men, transgender women

## Abstract

Mitigating HIV-related stigma is critical to improving HIV outcomes and reaching global HIV targets. We conducted formative community-engaged research to identify key characteristics of a social media campaign to address HIV-related stigma among youth in Lima, Peru. Focus groups and in-depth interviews with young people living with HIV (men who have sex with men, transgender women, sex workers, persons with perinatally-acquired HIV, cisgender women, and Venezuelan migrants), HIV advocates, and healthcare providers were carried out from November 2022 to July 2023. Audio recordings were transcribed verbatim and analysed for emergent themes using framework analysis. Two change targets were identified for campaign messaging: 1) changing how HIV and people living with HIV (PLWH) are perceived, and 2) changing attitudes and actions towards PLWH. Messages aligning with the first change target included education to raise awareness that HIV does not discriminate; HIV is a chronic, treatable condition; and “undetectable equals untransmittable” (U=U). Messages aligning with the second target included normalizing open conversations about HIV, encouraging support versus pity for PLWH, fostering unity among HIV-affected communities, and promoting inclusion of PLWH. Participants provided recommendations to achieve these messaging objectives, including how, to whom and by whom messages are delivered.

## INTRODUCTION

Stigma—defined as the social-cognitive process leading to the stereotyping and subsequent discrimination of individuals or groups[1]—poses an enormous threat to global health and impedes the control and treatment of infectious diseases such as human immunodeficiency virus (HIV). HIV-related stigma is associated with harmful consequences for people living with HIV (PLWH), including delayed diagnosis, treatment nonadherence, and nondisclosure to sexual partners[2-6]. Stigmatizing behaviours towards PLWH can manifest as loss of employment or housing and rejection by friends, family members and partners[7–9]. Additionally, HIV-related stigma is harmful to holders of stigmatizing beliefs, as it impedes the acceptability of preventative practices and accurate assessment of one’s own HIV risk[10-12].

There is promising preliminary evidence on the positive effects of anti-stigma campaigns and the potential for mass media communication strategies to normalize HIV[13-16]. Mass media campaigns (i.e., via television, radio, and/or newspapers) have led to widespread health-related behaviour changes[16], and those targeting HIV-related stigma have shown a modest impact on stigma reduction[15]. However, the elevated cost and decreasing popularity of these media limits their utility for stigma reduction. The global popularity of social media platforms, along with access to internet and mobile devices, allow for novel approaches to stigma reduction, whereby content can be widely and inexpensively disseminated. Social media is increasingly used as a platform for health communication campaigns, particularly targeting sexually transmitted infections[17-21]. While some of these interventions aim to reduce stigma, such as the *Get Yourself Tested* campaign in the United States[18], only one[19] included formal measures of stigma. This intervention successfully reduced stigma, but did not report community engagement to inform campaign content.

HIV-related stigma is common in Latin America[22], where there is additional pervasive stigma against populations in which HIV is mainly concentrated: gay, bisexual, and other men who have sex with men (GBMSM), transgender women (TGW), and cisgender female sex workers (FSWs)[23-29]. Recognizing the critical need to reduce HIV-related stigma and the relatively unexplored potential of social media platforms to reach both targets of stigmatizing behaviours and holders of stigmatizing beliefs, we conducted formative qualitative research with PLWH, HIV advocates, and HIV healthcare providers to inform an HIV stigma reduction social media campaign targeted to young people in Lima, Peru. We report findings on key messages, content delivery strategies, and overarching change goals for the campaign.

## METHODS

### Study setting

This work was conducted in Lima, Peru, where the majority of PLWH in Peru, an estimated 110,000, reside [29] and where a rising number of new HIV diagnoses in youth 18-29 has been reported[30]. HIV prevalence in Peru is estimated to be 0.3-0.4%, but is notably higher in transgender women (32%), GBMSM (10%), and FSWs (2.4%)[29],[31]. One recent study also found an elevated prevalence (1%) in Venezuelan migrants[32].

### Participant recruitment and consent process

From November 2022 to July 2023, we conducted focus groups and in-depth interviews with 110 participants living in Lima, Peru. Participants included a diverse group of young people living with HIV (YPLWH) (N=75) ages 16-29, recruited at health facilities and through referrals by healthcare providers and community partners. Participating YPLWH included GBMSM, TGW, FSWs, individuals with perinatal transmission, cisgender women, and Venezuelan migrants (Table 1). Additional sessions were conducted with healthcare providers (N=11), 73% of whom were women, and HIV advocates (N=24), 21% of whom were assigned female sex at birth and 25% of whom were TGW.

**Table 1:** Baseline characteristics of young people living with HIV (N=75)

| Characteristic | n (%) |
| --- | --- |
| Sex assigned at birth |  |
| Female | 26 (34.7) |
| Male | 47 (62.7) |
| Intersex | 2 (2.7) |
| Gender identity |  |
| Woman | 31 (41.3) |
| Man | 33 (44.0) |
| Transgender woman | 9 (12.0) |
| Genderfluid | 1 (1.3) |
| Other | 1 (1.3) |
| Sexual orientation |  |
| Homosexual | 29 (38.7) |
| Bisexual | 10 (13.3) |
| Heterosexual | 32 (42.7) |
| Other | 4 (5.3) |
| Median age, years (IQR) | 24 (21-27) |
| Sex worker | 3 (4.0) |
| Venezuelan migrant | 8 (10.7) |
| Perinatal HIV transmission | 17 (22.7) |

### Data collection

MW, a Peruvian qualitative researcher and certified nurse with experience working with YPLWH, conducted 13 focus groups and 16 in-depth interviews (the point of data saturation) using semi-structured interview guides (Supplemental Table 1) tailored to each participant group. These guides, informed by the Health Stigma and Discrimination Framework[33], were designed to facilitate discussion on experiences with HIV-related stigma, common myths associated with HIV, messages to be portrayed, and desired outcomes of the campaign. Sessions lasted between 60 and 120 minutes. In most sessions, only MW and participants were present; however, a team member aided group facilitation in five. Participants were compensated 50 soles (13 USD), plus any transportation costs. Focus groups with healthcare personnel and all interviews were conducted virtually. Notes were not taken. No repeat interviews were conducted. Sessions were audio-recorded, transcribed verbatim, and translated from Spanish to English. Transcripts were not returned to participants for comment or correction.

### Data analysis

Transcripts were uploaded into the qualitative research software Dedoose. Two qualitative analysts (AN, MW) conducted a systematic, comparative, and inductive analysis of the transcripts guided by the Framework Analysis[34]. Initial codebooks were guided by the interview guide and Health Stigma and Discrimination Framework[33]; additional codes were created *de novo* as needed. Both analysts reviewed each transcript to reach a consensus on codes, which were organized into emergent themes. Here, we report results from excerpts related to change targets and messages for achieving these goals. The COREQ checklist was used for rigor and transparency[35].

### Research ethics

The Peruvian Vía Libre Bioethics Committee (Approval 8326) and Harvard Medical School Institutional Review Board (Approval IRB22-0741) approved this study. Participants and guardians of participants <18 years old provided written informed consent or assent, as applicable. The study was granted a waiver of consent for adolescents <18 years old who had not disclosed their HIV status to a guardian.

## RESULTS

Among messages that participants identified as fundamental components of an anti-stigma campaign, two categories emerged as targets (i.e., change goals): (1) messages that change perceptions of HIV and PLWH, and (2) messages that change attitudes and behaviours towards PLWH. Within these, subcategories of educational messaging were proposed to challenge the stereotypes and prejudices that lead to discrimination.

### Messages that change perceptions of HIV and PLWH

#### HIV does not discriminate

Participants highlighted the need to communicate that anyone is at risk of contracting HIV regardless of sexual orientation, gender identity, age, or social class, as HIV *“doesn’t distinguish social condition. It doesn’t matter if you’re a housewife or if you’re faithful, or if you’re not trans, or not gay. All the same, you can [still have HIV]*.*”* (YPLWH, cisgender woman) To combat the widespread belief that HIV only affects specific populations, specifically the LGBTQIA+ community, participants emphasized that messaging should clarify that risk is connected to behaviours (i.e. condomless sex) and exposures rather than to groups. Participants agreed that an effective campaign would explain these behaviours within a broader conversation about modes of transmission and address how HIV is not transmitted to counter misconceptions that sharing utensils or having non-sexual physical contact with PLWH implies a risk of acquisition. One healthcare provider emphasized:

> *The majority of people think that because they use the same towel [*…*] or use the same soap, they’re going to, or someone’s going to give them HIV. Obviously, we know it’s not like that, but it would be great to make it clear that it’s through sex, or from needles, or transmission from mother to child*. (Infectious Disease Specialist)

#### HIV is a treatable chronic condition

Participants recommended the campaign clearly present HIV as a treatable chronic condition to change the narrative that an HIV diagnosis is a certain death sentence. One participant explained, *“people should view it as if it was cancer or diabetes [*…*], something like that. Because people see [HIV] as something so bad*.*”* (YPLWH, FSW) They emphasized that with proper adherence to freely available treatment, PLWH can live normal, healthy lives, studying, working, and having families. Participants expressed that these messages should also reach young people receiving a new HIV diagnosis. One participant stated:

> *For someone who’s just been recently diagnosed, one of the messages should be that your life’s going to go on like always. Because a lot of people think their life is done, it’s over, or they’re not going to be able to work, or they’re going to be rejected from jobs, things like that. The first important message would be for them to understand that their life’s going to go on like it always has. It’s not going to change. It’s just a condition that’s going to be there*. (YPLWH, GBMSM)

Participants advocated for messages describing the differences between HIV and AIDS to counter the perception that the two are synonymous and associations of HIV with extreme sickness and death. Instead, campaign messages should focus on living, vitality and the right for PLWH to participate in all aspects of society without stigmatization.

#### Undetectable = Untransmittable

Participants spoke of the need to educate the general public that U = U, meaning that PLWH who achieve an undetectable viral load through adherence to antiretroviral treatment (ART), do not transmit HIV to others through sex. Many described their peers as not fully understanding the implications of this statement, *“*… *they think there’s a but. So, I’m undetectable, but maybe there’s a risk [*…*]Even though there have been many campaigns, people are still scared of transmission, even if you’re undetectable*.*”* (YPLWH, GBMSM) Participants felt messages should explain the effectiveness of ART and clarify that it is possible for PLWH to be sexually active without risk of transmitting HIV.

Additional illustrative quotes are included for each subcategory in Table 2.

**Table 2.** Changing perceptions of HIV and PLWH: Subcategories, messages and illustrative quotes.

| *Subcategory* | *Messages* | *Illustrative quotes* |
| --- | --- | --- |
| HIV does not discriminate. | a. Anyone can contract HIV.<br>b. HIV is a condition of behaviours and exposures rather than populations.<br>c. How HIV is and is not transmitted | *That people behind the screen watch this and say "wow [...] it's not just such and such people who acquire it, it could also happen to my aunt, to my dad who's a doctor, to my brother who's a student." (YPLWH, FSW)*<br>*That HIV is a condition of behaviours, not of groups. That anyone can acquire HIV, not just those vulnerable groups, which is what we've been taught for so long. (Infectious Disease Specialist)*<br>*Sharing, eating, drinking together, at a meet up, having fun together, we can't transmit [HIV] to them that way. That they know that transmission is only through blood, through sex, only that. (YPLWH, cisgender woman)* |
| HIV is a treatable chronic condition. | a. HIV is not a death sentence.<br>b. HIV is not AIDS.<br>c. PLWH can lead lives similar to those without HIV. | *Make it clear that in this day and age, HIV isn't synonymous with death. What's more, people living with HIV have a normal life. Their life expectancy is the same as someone not living with HIV. Also, their quality of life is the same as someone not living with HIV. [Infectious Disease Specialist]*<br>*And reinforce that we don't have AIDS, that AIDS is the last stage because a lot of times straight friends come and say, "Oh, you have AIDS, I'm sorry, you poor thing, how much time do you have left?" (HIV advocate)*<br>*That people know that just because you're living with HIV doesn't mean your whole world ended [...] because someone living with HIV can live a normal life, you know, that would be the most important. (YPLWH, TGW)* |
| Undetectable = Untransmittable. | a. PLWH can have a fulfilling sex life without risk of transmission to their partners.<br>b. PLWH can start a family and have children without putting them at risk of HIV. | *People should have information and knowledge, and they should know that people who are undetectable don't transmit [HIV]. (YPLWH, TGW)*<br>*Normalize the topic that if you have HIV you can still have sex. There are a lot of people I see who think that because they have HIV they can no longer have sex. (HIV advocate)*<br>*I would like everyone to be clear that those of us who have HIV are taking medication, we have a life like anyone else's, with the same conditions. Basically that, you know? That yes, we are taking medication, that yes, we can be ok, we can start a family. (YPLWH, perinatal transmission)* |

### Messages that change attitudes and behaviours towards PLWH

#### Normalize talking about HIV

Participants hoped that the anti-stigma campaign would lead to greater freedom to discuss an HIV diagnosis without “… *that fear, that I don’t want to talk about HIV precisely because I could be rejected, but rather that we’re free to talk about it”* (YPLWH, perinatal transmission) and without *“*… *the fear that if someone finds out, what are they going to think*.*”* (YPLWH, perinatal transmission) Participants felt that to achieve this, campaign content should present HIV as a normal topic of conversation among family and friends and demonstrate how taboos around discussing HIV and resulting discrimination negatively affect PLWH.

In participant experience, when HIV emerges in conversation, it is frequently accompanied by discriminatory remarks or simple indifference. As one participant noted, to counter these reactions the campaign should “*show what those words do to people, to us, how they affect our lives*.*”* (YPLWH, GBMSM)

#### Encourage support instead of pity for PLWH

Participants wanted campaign messages to increase awareness of what it means to live with HIV so as to remove feelings of pity. As one participant described, “*I don’t want people to feel bad for me [*…*] and I also don’t want to hear ‘oh you poor thing’, because I’m not a poor thing. I’m not a poor thing because I live with this*.” (YPLWH, FSW) Participants living with HIV instead seek support, encouragement, and equal treatment from society. Thus, it was suggested that messages educate audiences on how to approach HIV with empathy and also address feelings of self-stigma and guilt among PLWH, particularly those with new diagnoses.

Though participants hope the campaign will lessen judgment of PLWH by society as a whole, special mention was made of messages targeting stigma by healthcare providers. Participants suggested messages foster reflection on how explicit and implicit acts of discrimination at health facilities could drive PLWH away from medical care and on how all populations living with HIV deserve equal care, highlighting that “*the health system has a responsibility to treat us with the same warmth and quality that everyone deserves*.” (HIV advocate)

#### Promote inclusion

Participants living with HIV frequently cited a desire to be accepted and included in society just as anyone else. They suggested campaign messages present acceptance of PLWH as a step towards inclusion, as many times people “*know that there are people living with HIV, but that’s all, at a distance, yeah I support them, I accept them no problem, but over there, leave them over there*.” (YPLWH, GBMSM, Venezuelan migrant) However, they highlighted that true inclusion should go beyond acceptance to the active invitation to participate in all aspects of society. Participants recommended that messages challenge the public to include PLWH in their daily lives, especially in the workplace where participants mentioned facing particular stigma and exclusion. One participant described, “*I was working in a warehouse, they made me do a blood test and it came back positive [for HIV]. I had been working fine, but it was really uncomfortable to have gone through the whole process and then they say ‘no you can’t work here because you have this*.’” (YPLWH, GBMSM)

#### Foster unity among HIV-affected communities

Several participants discussed their perceptions that the LGBTQIA+ community itself is fractured and that for the general public to accept and include the community, first “… *the community itself has to heal, because if we don’t make a change, how can we expect other people to accept us and treat us differently*?” (YPLWH, GBMSM, Venezuelan migrant) They recommend messages should strive to break down the stereotypes that different groups affected by HIV have of one another to promote a united front against HIV-related stigma. As one participant noted, “*the [LGBTQIA+] community has grown a lot [*…*]But it’s time we break down all these stereotypes because we’re all one community and we’re all susceptible to HIV, to stigma, and to all the prejudice that exists*.*”* (HIV advocate)

Additional illustrative quotes are included for each subcategory in Table 3

**Table 3.**
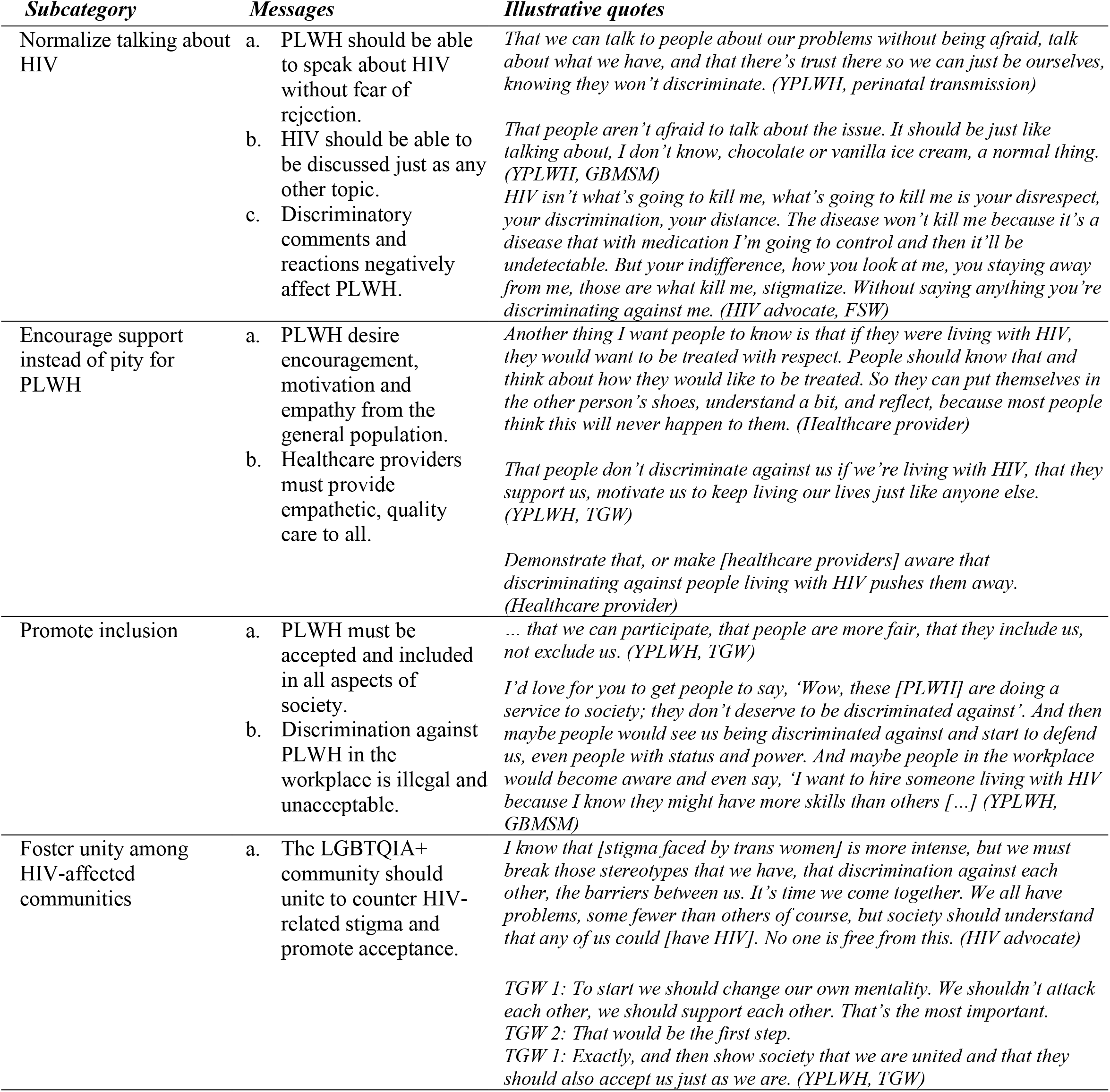
Changing attitudes and behaviours towards PLWH: Subcategories, messages and illustrative quotes.

| *Subcategory* | *Messages* | *Illustrative quotes* |
| --- | --- | --- |
| Normalize talking about HIV | a. PLWH should be able to speak about HIV without fear of rejection.<br>b. HIV should be able to be discussed just as any other topic.<br>c. Discriminatory comments and reactions negatively affect PLWH. | *That we can talk to people about our problems without being afraid, talk about what we have, and that there's trust there so we can just be ourselves, knowing they won't discriminate. (YPLWH, perinatal transmission)*<br>*That people aren't afraid to talk about the issue. It should be just like talking about, I don't know, chocolate or vanilla ice cream, a normal thing. (YPLWH, GBMSM)*<br>*HIV isn't what's going to kill me, what's going to kill me is your disrespect, your discrimination, your distance. The disease won't kill me because it's a disease that with medication I'm going to control and then it'll be undetectable. But your indifference, how you look at me, you staying away from me, those are what kill me, stigmatize. Without saying anything you're discriminating against me. (HIV advocate, FSW)* |
| Encourage support instead of pity for PLWH | a. PLWH desire encouragement, motivation and empathy from the general population.<br>b. Healthcare providers must provide empathetic, quality care to all. | *Another thing I want people to know is that if they were living with HIV, they would want to be treated with respect. People should know that and think about how they would like to be treated. So they can put themselves in the other person's shoes, understand a bit, and reflect, because most people think this will never happen to them. (Healthcare provider)*<br>*That people don't discriminate against us if we're living with HIV, that they support us, motivate us to keep living our lives just like anyone else. (YPLWH, TGW)*<br>*Demonstrate that, or make [healthcare providers] aware that discriminating against people living with HIV pushes them away. (Healthcare provider)* |
| Promote inclusion | a. PLWH must be accepted and included in all aspects of society.<br>b. Discrimination against PLWH in the workplace is illegal and unacceptable. | *... that we can participate, that people are more fair, that they include us, not exclude us. (YPLWH, TGW)*<br>*I'd love for you to get people to say, 'Wow, these [PLWH] are doing a service to society; they don't deserve to be discriminated against'. And then maybe people would see us being discriminated against and start to defend us, even people with status and power. And maybe people in the workplace would become aware and even say, 'I want to hire someone living with HIV because I know they might have more skills than others [...]' (YPLWH, GBMSM)* |
| Foster unity among HIV-affected communities | a. The LGBTQIA+ community should unite to counter HIV-related stigma and promote acceptance. | *I know that [stigma faced by trans women] is more intense, but we must break those stereotypes that we have, that discrimination against each other, the barriers between us. It's time we come together. We all have problems, some fewer than others of course, but society should understand that any of us could [have HIV]. No one is free from this. (HIV advocate)*<br>*TGW 1: To start we should change our own mentality. We shouldn't attack each other, we should support each other. That's the most important.*<br>*TGW 2: That would be the first step.*<br>*TGW 1: Exactly, and then show society that we are united and that they should also accept us just as we are. (YPLWH, TGW)* |

The underlying theme of all messages was that PLWH are no different from anyone else, with similar hopes, fears, and aspirations, and should not be seen *“*… *as good or bad, but rather just as normal people, like anyone else, with the same life expectancy, the same access, the same ability to work, the same difficulties as anyone else*.*”* (HIV advocate)

### Content Delivery

#### Videos

Participants agreed that short videos (15-30 seconds) with captivating intros are the most powerful means of conveying anti-stigma messages. The following types of videos were mentioned as potentially most effective: storytimes (a TikTok trend in which someone films themselves telling a story in a casual setting often while doing another activity), testimonials both positive and negative, and social experiments in which PLWH interact with the public. Participants considered that spokespeople in these videos should represent diverse identities in all aspects and especially gender, sexual orientation, race, socioeconomic status, education, and religion. As one participant advised, “*Pay attention to the profiles you choose. They should be varied in terms of orientation, identity, race, income, and everything else, because it’s crosscutting, you know*.*” (HIV advocate)*

Generally, participants felt that PLWH would be more credible than actors sharing these messages and that it would be essential that all actors and those creating content or disseminating the campaign receive training on HIV and HIV-related stigma. Most agreed that “*the majority should be PLWH, but also show people who aren’t living with HIV because maybe they’re also affected and want to talk about it*.*” (HIV advocate)* According to participants, family members, friends, or colleagues of PLWH would be compelling messengers of anti-stigma information.

> *Focus group leader: Who should give these messages?*
>
> *TGW 1: A trans woman, a gay man, an actor, or whoever you think is convenient and would do a good job*.
>
> *TGW 2: Exactly. To get a clear and direct message to the public. [YPLWH, TGW]*

#### Audience

Many participants, particularly those involved in HIV activism, noted that the majority of media campaigns related to HIV are aimed at the LGBTQIA+ community. They agreed that this reinforces the stereotype that HIV is only an issue for a single population and suggested that an anti-stigma campaign should be aimed at all audiences equally.

> *Also don’t just shut yourself in the community, I mean the gay and trans community, it would also be good for the general population to be informed. Because the campaigns they do, it’s true, they’re just for the gay and trans community, key populations. The general population always asks, always has questions, so to break with that misinformation they have, it would be great to share this with everyone. (HIV advocate)*

#### Language

The importance of careful selection of language was another key area related to content delivery. In Spanish, participants preferred ‘transmitir’ (to transmit) over ‘contagiar’ (to infect) when discussing HIV-acquisition, and ‘vivir con VIH’ (to live with HIV) or ‘ser seropositivo’ (to be seropositive) to identify PLWH. Participants described a feeling of coexisting with the virus as *“[HIV] is just one of the many qualities one may have” (HIV advocate)* and as such agreed that HIV should be described as a health or life condition rather than a disease.

#### Images

The red ribbon symbolic of HIV was controversial when discussing imagery to associate with an HIV stigma campaign. Many participants felt that the red ribbon is in itself stigmatizing as it labels the user as having some relationship with AIDS. Others mentioned that the red ribbon has been over-used and that ribbons, in general, are employed in sad or negative situations, contradicting the campaign’s objective of eliminating feelings of pity towards PLWH. However, a sub-group of activist participants noted that, for them, the red ribbon is a symbol of solidarity.

Participants suggested not using images frequently associated with HIV-campaigns, such as condoms, pills, or drops of blood, and were in favour of a unique logo, perhaps one incorporating a subtle reference to the red ribbon (e.g. an animal mascot wearing a red scarf). In general, “*[Imagery should be] as normal as possible and not related to disease. If there are condoms or pills, then you are referencing the disease*.*” (YPLWH, GBMSM)* The use of the LGBTQIA+ flag in the campaign was met with caution as it could perpetuate the stereotype that HIV only affects members of this community.

Supplemental Table 2 includes additional illustrative quotes of campaign language and imagery.

## DISCUSSION

In this study, a diverse group of YPLWH and other stakeholders, including healthcare providers and advocates, identified two overarching targets (i.e., change goals) for a campaign to reduce HIV-related stigma in Peru: to change how PLWH are perceived and to change attitudes and actions towards PLWH. While these change targets emerged organically, they coincide with theoretical models for stigma and stigma reduction that posit that a lack of correct information leads to prejudices and stereotypes and, ultimately, discrimination (also known as enacted or experienced stigma), and that stigma change programs should include specific change targets or goals(1)[33]. The messaging associated with the two change goals, while initially targeted broadly toward youth living in Peru, also has the potential to positively impact YPLWH by reducing self-stigma (i.e., stigma that results when a member of a stigmatized group believes the stereotypes and prejudices associated with the stigmatized condition and applies them to the self). In Peru, like many places in Latin America, PLWH frequently experience a high level of self-stigma[28].

Results suggest that messaging aligned with the first change target should convey factual information about HIV and what it means to live with HIV. Three specific messages were identified as critical: (1) HIV does not discriminate (i.e., no one is immune from potentially acquiring HIV); (2) HIV is a treatable chronic condition; and, (3) if HIV is controlled (i.e., undetectable) in one’s body, it will not be transmitted sexually (i.e., U=U, or I=I in Spanish). Providing education to correct common misconceptions and erroneous beliefs (e.g., “HIV is synonymous with death”, “PLWH cannot have sex or start a family without risking HIV transmission to their partner or baby”) is a destigmatizing strategy that can reduce the prejudices and stereotypes that lead to discriminatory behaviour[36]. The potential for community-derived U=U messaging in combatting stigma reduction has emerged as a particularly promising educational messaging strategy[37],[38]; however, less awareness of this concept has been reported in low- and middle-income countries[38]. Messages aligning with the second change target aimed to address prejudice and discrimination through actions, including normalizing open discussion about HIV, offering support versus pity, ensuring inclusion in all aspects of societal life, and fostering unity within the different communities most affected by HIV. Much attention has been paid to reducing stigma toward members of stigmatized groups, but less to fostering affirming attitudes and building allyships, which is critical to improving outcomes for stigmatized groups[39].

Translating key messages to content appealing to young people is critical to ensuring effectiveness. Our findings offer practical suggestions for the creation of content to communicate these messages, some of which broadly align with prior studies[39-41]. Among these is the importance of diversity in representation to ensure relatability across various identities and because a focus on a single population, such as the LGBTQIA+ community, may reinforce existing stigmas[40],[41]. We found that content imagery and tone are also key considerations. Imagery that reinforces stereotypes or that is overly negative or aggressive may be less effective than lighter messages, free of shame and judgment[42]. For example, several participants referenced antiquated images from the pre-ART era that immortalize the misconception that HIV is analogous to death. An upbeat tone with positive imagery may more appropriately reflect the lifesaving advances in HIV treatment. Other research offers pragmatic guidance for social media strategies targeting young people. For example, one study found that provocative images and wording to grab the user’s attention, followed by fact-based content, were effective in motivating engagement with services offline[42].

Language emerged as another important consideration. Although a desire to avoid stigmatizing words is intuitive, language’s subjective and evolving nature may challenge this goal. Some participants noted that they preferred the term “seropositive” to describe themselves and others living with HIV, while other stakeholders in our study and beyond have identified the term as stigmatizing[43],[44]. Community advisory committees, ideally comprised of people of similar ages as the target population, should play a key role in validating campaign content and ensuring that the language is correct, understandable, acceptable, and appealing without reinforcing stigma.

A strength of this work was the engagement of a diverse group of YPLWH representative of the groups most impacted by HIV in Peru. Effective community engagement to inform campaign messaging ensures that campaign priorities align with those of the most affected groups. While qualitative methodologies are one strategy to elicit community input[41], other community-based participatory methods such as crowdsourcing[37],[41],[44], and discrete choice experiments have also been used to engage communities in public health campaigns. By focusing on the opinions of PLWH and incorporating those of health providers and advocates, we aimed to elevate the voices of those with lived experience. We did not validate our findings among the potential targets of the campaign (i.e., young people without a known HIV diagnosis). If there were a misalignment between the messaging strategies identified by study participants and those of the campaign targets, this could undermine the effectiveness of subsequent campaign content. Finally, although we aimed to explore messages about stigmas that intersect with HIV (i.e., stigmas against GBMSM or TGW), this proved challenging. While participants readily shared experiences of stigma due to different parts of their identity, we could not generate in-depth discussion around lived experiences at the intersection of these stigmas. Intersectional stigma is conceptually difficult to measure, perhaps even more so in young people with fewer years of lived experience. Our qualitative guides likely required greater depth and attention to the lived experiences of intersectional stigmas than we gave them in this multi-aim study[45].

## CONCLUSIONS

We employed a community-engaged approach to identify change goals, messages, and messaging preferences to underpin a Spanish-language social media campaign targeting young people. While this formative research was exclusive to Peru, the findings broadly align with the existing literature and likely have broad applicability, expanding the evidence base regarding the use of social media to reduce stigma related to HIV and, potentially, to other stigmatized conditions.

## ACKNOWLEDGEMENTS

The authors are indebted to the study participants, and to the healthcare providers and community partners who supported the study team in participant recruitment. We would also like to express our gratitude to the Fogarty International Center for their financial support and to the Socios en Salud Youth Advisory Board for their guidance throughout our project.

## AUTHORS’ CONTRIBUTIONS

Study design: M.W., K.K., J.G., M.F., R.E.

Analysis design: K.K., J.G., M.F., R.E.

Study implementation: M.W.

Data management: A.N., M.C., M.W.

Data analyses: A.N., M.C., M.W., K.K., R.E.

Data interpretation: all authors

First draft: A.N., M.C., K.K, M.F., R.E.

Critical review of manuscript: all authors

Approved final version: all authors

## DATA AVAILABILITY

The data that support the findings of this study are available upon reasonable request from the corresponding author. The data are not publicly available due to privacy or ethical restrictions.

