## Supplemental Table 1 for "Change targets, messaging, and content delivery for a community-engaged social media campaign addressing HIV-related stigma in Peru: a qualitative study"

**Supplemental Table 1. Qualitative data collection guide developed for focus groups and interviews with PLWH**

| Domain | Question | Probes |
|---|---|---|
| **Understand experiences with HIV-related stigma, including various types of intersectional stigma** | • How would you describe “stigma”?<br>• Tell me about your experience with stigma and HIV.<br>• Tell me about your experience with stigma and other parts of your identity?<br>• How does your identity as [X] relate to your experiences of HIV and stigma?<br>• From whom or which type of person do you most experience (or have a fear of) being stigmatized?<br>• What area(s) of your life are affected by HIV stigma?<br>• By whom do you feel the least stigmatized or most valued/empowered? | • What is stigma?<br>• How has stigma affected you?<br>• Have you been affected by other types of stigma (i.e., other than HIV)?<br>• What additional challenges do you face due to living with HIV and having the identity of [X]?<br>• What effect does this stigma have on you (what you can/cannot do, say, etc.)?<br>• Family? Friendships? Relationships? School? Church? Health care? Work?<br>○ Which is the most strongly impacted?<br>• Why do you think this is? |
| **Define key intervention messages** | • What are the most common stereotypes or misconceptions that others hold about HIV?<br>• As a person living with HIV, how would you like to be perceived by others?<br>• What message(s) would you most want other people to know about HIV?<br>• Imagine you were creating social media posts with anti-HIV stigma messages. What messages would you show?<br>• Imagine you were creating social media posts with anti-HIV stigma messages. What images would you show? | • How have you personally experienced or seen these?<br>○ Which of these impacts you the most? Why?<br>• Can you list some adjectives?<br>• How do these messages differ by the type of person (for example, family or friend or doctor or teacher)?<br>• You could use pictures, memes or videos. Be creative!<br>• Can you think of images, words, or symbols that represent you or that represent an anti-stigma message? |
| **Identify change goals** | • What is the one thing that you would like to change from a stigma reduction intervention?<br>• What is the one thing that you would most like to change regarding how people living with HIV are treated? | • Why?<br>• How would your life be different if this were to come true? |
| **Identify influencers not previously considered** | • Here is a list of well-known social media influencers we plan to work with. What are your opinions about this list?<br>• On social media, whose opinions do you value? | • Do you know them? How?<br>• Anyone you would remove?<br>• Anyone you would add?<br>• From what types of people do you value information?<br>• Can you provide some specific examples? |
| **Information on content** | • When you are scrolling through your feed, what makes you stop at a post?<br>• What types of social media content do you enjoy most?<br>• Are there words that make you know the content is geared toward young people like you?<br>• Should there be a project mascot? Spokesperson? If so, what should they be like? | • What form does it take? (video, cartoons, drawings, music)<br>• Memes? Tiktoks? Videos? Narratives/stories? News updates? Informational?<br>• List as many examples as you can.<br>• Funny? Serious? Everyday? Attractive? Living with HIV? |
