## Supplemental Table 2 for "Change targets, messaging, and content delivery for a community-engaged social media campaign addressing HIV-related stigma in Peru: a qualitative study"

**Supplemental Table 2. Illustrative quotes for campaign language and imagery**

| |
|---|
| ***Language*** |
| *It feels much more freeing to me to say I'm living with HIV. [PLWH, GBMSM]* |
| *I prefer the term seropositive. [PLWH, GBMSM]* |
| *HIV isn't a "disease", it's a lifelong condition. [HIV advocate]* |
| ***Imagery*** |
| *Lately the ribbon is being used for sad occasions, something that symbolizes someone that's died, really the idea should be something that associates HIV with life. [HIV advocate]* |
| *No, the red ribbbon, it's been overused. [PLWH, heterosexual]* |
| *To us the ribbon is solidarity, it's not disease. It's not death. It's solidarity. [HIV advocate]* |
